# Changes in health and functioning of care home residents over two decades: what can we learn from population based studies?

**DOI:** 10.1101/2020.08.05.20168740

**Authors:** Robert O Barker, Barbara Hanratty, Andrew Kingston, Sheena Ramsay, Fiona E Matthews

## Abstract

**Background:** Care home residents have complex care and support needs, as demonstrated by their vulnerability during the COVID-19 pandemic. There is a perception that the needs of residents have increased, but evidence is limited. We investigated changes in health and functioning of care home residents over two decades in England and Wales.

**Methods:** We conducted a repeated cross-sectional analysis over a 24-year period (1992-2016), using data from three longitudinal studies, the Cognitive Function and Ageing Studies (CFAS) I and II and English Longitudinal Study of Ageing (ELSA). To adjust for ageing of respondents over time results are presented for the 75-84 age group.

**Results:** Analysis of 2,280 observations from 1,745 care home residents demonstrated increases in severe disability (difficulty in at least two from washing, dressing and toileting). The prevalence of severe disability increased from 63% in 1992 to 87% in 2014 (subsequent fall in 2016 although wide confidence intervals). The prevalence of complex multimorbidity (problems in at least three out of six body systems) increased within studies over time, from 33% to 54% in CFAS I/II between 1992 and 2012, and 26% to 54% in ELSA between 2006 and 2016.

**Conclusion:** Over two decades, there has been an increase in disability and the complexity of health problems amongst care home residents in England and Wales. A rise in support needs for residents places increasing demands on care home staff and health professionals. This is an important concern for policymakers when considering the impact of COVID-19 infection in care homes.

**Key points:**

- Care home residents have complex care and support needs, which has contributed to their vulnerability during the COVID-19 pandemic.
- Despite a perception that the needs of care home residents have increased over time, the epidemiological evidence is limited.
- This study demonstrates an increase in the level of disability and the complexity of health problems amongst care home residents in England and Wales over two decades.
- The rise in support needs for care home residents places increasing demands on care home staff and health professionals, and should be an important consideration for policymakers and service commissioners when considering the impact of current and future waves of COVID-19 infection in this setting.

## Background

Care home residents are known to have high needs for health and social care support [1]. There is a perception amongst care home and NHS staff that residents' needs have grown in number and complexity in recent years [1,2]. However, evidence from epidemiological studies about changes in the resident population is limited. People in care homes take part in some research, but many studies exclude residents, either at the start, or at the point when they move into a care home [3].

Information about UK care home residents is available from a small number of population-based cohort studies, including the Cognitive Function and Ageing Studies (MRC CFAS – here called CFAS I and CFAS II)[4], and the English Longitudinal Study of Ageing (ELSA) [5]. Analysis of these and other data point to a possible rise in care needs. A study based on ELSA (2002 to 2015) described an increase in the number of health conditions and functional deficits amongst older people who were about to move into a care home [6]. The proportion of older people living with dementia in care settings increased from 56% in CFAS I (1991-1994) to 70% in CFAS II (2004) [7]. Despite nearly all residents in long-term care having functional impairment at both time points, more were chair or bed-bound in CFAS II (34%) compared to CFAS I (22%) [8].

An in-depth understanding of trends in the health and functioning of care home residents is needed as care providers and policy makers strive to meet the needs of this complex population. The COVID-19 pandemic has highlighted the consequences of this gap in our understanding of the health of care home residents and the level of support required. In the absence of a minimum dataset on UK care home residents, this study set out to synthesise data from existing cohort studies of ageing in England in Wales. The aim is to investigate how the health and functioning of care home residents in epidemiological studies in England and Wales has changed over time. This will be done by addressing the question of how the proportion of care home residents experiencing complex multimorbidity, severe disability and poor self-reported health has changed over time.

## Methods

### Study design and participants

Data were obtained from MRC-CFAS (CFAS I), CFAS II and ELSA. Full details of the CFAS I [9] and CFAS II study design and methods have been described in detail elsewhere [7]. Briefly, CFAS I and II are both interview population-based cohort studies [8]. The CFAS I interviews were conducted between 1991 and 2003 in five geographical areas in the UK (Cambridgeshire, Gwynedd, Newcastle, Nottingham, Oxford). CFAS II interviews took place between 2008 and 2012 using three of the same geographical areas (Cambridgeshire, Newcastle, Nottingham) and the same study design. ELSA is a panel study of men and women aged ≥ 50 years living in England [5]. Participants are interviewed approximately every 2 years [5]. ELSA does not recruit new participants from care homes, but attempts to collect data on those who transfer into a care home during the follow-up period [6]. The ELSA core datasets for each individual wave were used to identify all participants who had an institutional interview, either in-person or informant, to define the care home population. The ELSA harmonized dataset, which incorporates proxy responses, was then used to investigate the variables of interest. Individuals were included if they were original participants, or refreshment sample participants (partners were excluded) for all interviews from wave 3 to wave 8. In addition, to reflect the ageing sample, individuals who were below the age of 65 at each interview wave were excluded. In CFAS I and II, participants living in council residential or nursing homes, private nursing homes or long stay hospitals defined the care home population. CFAS I and II used informant interviews to supplement respondent information in the case of cognitive or physical frailty impairing the interview. Respondent and interview information was merged.

### Variable description

We explored changes in self-reported health, levels of comorbidity and disability. Self-reported health was reported as excellent, good, fair or poor in ELSA and CFAS studies. For the multimorbidity and disability domains, we selected core variables that were common to CFAS and ELSA, to allow comparison across datasets. These core variables were combined into new variables, which were used to make inferences about multimorbidity and disability. The variable groupings are shown in tables 1 and 2 in the supplementary material.

**Table 1:** Data sources and participant characteristics.

| **Study** | **Round of data collection** | **Modal year** | **Number of participants in care homes.**<br>**All / First interview** | **Median age**<br>**All / First interview** | **Gender**<br>**(% female)** |
| --- | --- | --- | --- | --- | --- |
| CFAS I | SO | 1992 | 630 / 630 | 84 / 84 | 74.9 |
| CFAS I | S2, C2 | 1994 | 574 / 331 | 84 / 83 | 73.3 |
| CFAS I | S6, C6 | 1997 | 256 / 130 | 87 / 86 | 76.4 |
| CFAS I | CX | 2002 | 148 / 125 | 87 / 88 | 77.8 |
| ELSA | Wave 3 | 2006 | 47 / 47 | 85 / 85 | 72.2 |
| ELSA | Wave 4 | 2008 | 62 / 45 | 86 / 87 | 77.0 |
| ELSA | Wave 5 | 2010 | 69 / 47 | 86 / 86 | 71.3 |
| ELSA | Wave 6 | 2012 | 72 / 46 | 88 / 88 | 79.2 |
| ELSA | Wave 7 | 2014 | 59 / 34 | 88 / 88 | 68.8 |
| ELSA | Wave 8 | 2016 | 56 / 44 | 91 / 91 | 79.7 |
| CFAS II | Wave 1 | 2010 | 202 / 202 | 86 / 86 | 71.6 |
| CFAS II | Wave 2 | 2012 | 105 / 64 | 88 / 87 | 74.0 |

Disability was measured as difficulty in undertaking activities of daily living. Activities relevant to care home residents were included, such as help with going to the toilet. Variables less relevant to care home residents, higher on the hierarchy of disability [10], for example the ability to do shopping and prepare a meal, were excluded. Participants were classified as having severe disability if they had difficulty, or needed assistance, with two out of the three domains - washing, dressing and going to the toilet.

In the multimorbidity domain, we selected variables that have been shown to be risk factors for functional decline in older adults [11]. The medical conditions in the core variables were grouped according to body system: cardiovascular, cerebrovascular, musculoskeletal, endocrine, respiratory or cognition. The resident was classified as having comorbidity in a domain if they had a medical condition related to that particular body system. Complex multimorbidity was defined as having a comorbidity in at least three out of six body systems [12] (table 2, supplementary material).

### Statistical analysis

STATA15 was used to conduct a repeated cross-sectional analysis study. At the mean time point for each round of data collection, the prevalence of core and derived variables was calculated. In CFAS I and II, the interview question about medical conditions was sometimes phrased according to responses at a previous wave of data collection ('since we last saw, has your Doctor told you…'). In these instances, responses from previous rounds of data collection were considered in order to derive the prevalence. Baseline (cross sectional) weights were released into both studies. To adjust for longitudinal attrition inverse probability weighting was calculated for each wave taking account of age, sex, health status, disability, self-reported health, and care home status at previous wave. To account for item non-response within an interview, multiple imputation was undertaken. Each study had its own multiple imputation model for all variables that are used to calculate self-reported health, multimorbidity and severe disability, together with care home status, age and sex. Multiple imputation by chained equations using 50 imputation samples were used. Logistic regression was used to model the relationship between the presence of each variable with age and wave of interview. The main analysis presents predicted probabilities of each variable within the age group 75-84 years, which are used in the analysis to adjust for the longitudinal nature of the data and as small numbers make age-standardisation unstable. A sensitivity analysis to account for including the same individuals at more than one interview presents only the cross-sectional results from the individuals the first time they were interviewed in a care home (see supplementary appendix).

## Findings

### Study Participants

We analysed 2,280 observations from 1,745 care home residents of which 74% were female. The mean age was 85. Data sources and participant characteristics are presented in Table 1. Data were obtained from 365 observations from the ELSA cohort, waves 3 (2006) to 8 (2016). There were no participants in care homes in waves 1 or 2. There were 1,608 observations from care homes across four waves of CFAS I, spanning a period from 1992 to 2003. There were 307 observations across the two waves of data collection for CFAS II, from 2009 to 2013.

### Functioning and disability

There is an emerging trend showing that the prevalence of severe disability is increasing between 1992 (63% - CFAS) and 2014 (87% - ELSA) (Supplementary material: table 1 and figure 1). This trend is largely driven by increases in the prevalence of difficulty, or needing assistance with, dressing and bathing. A less clear pattern was observed for toileting. The prevalence of severe disability fell in 2016 (ELSA) though the estimate has a wide confidence interval and more data is needed to determine whether this fall persists, given the consistency of increases demonstrated in previous years.

**Figure 1:**
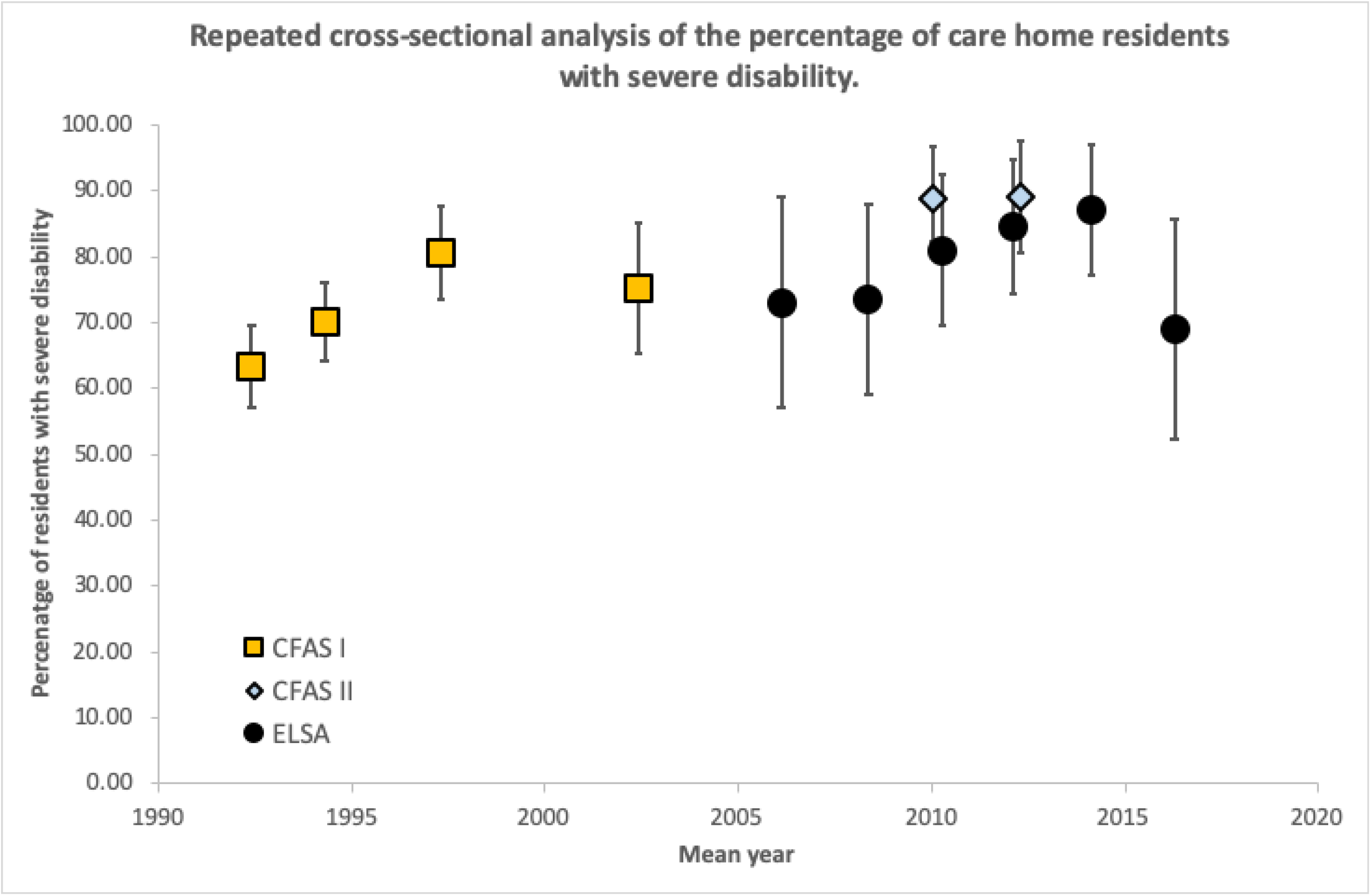
Prevalence of severe disability amongst care home residents from 1992 – 2016 in CFASI/II and ELSA studies. Severe disability defined by difficulty in two out of the three domains (washing, dressing, toileting).

### Health and multimorbidity

The prevalence of complex multimorbidity amongst care home residents showed increases within each study over time, from 33% to 54% in CFAS I/II (1992 to 2012) and from 26% to 54% in ELSA (2006 to 2016), but not overall (Figure 2, Supplementary Table 2). This may reflect the differences in multimorbidity reporting and the diagnostic criteria used for each study. For CFAS I and II, the prevalence of problems with cognition increased from 75% in 1992 to 95% in 2012. ELSA exhibited a similar trend, increasing from 60% in 2006 to 81% in 2016. Likewise, for CFAS I and II, the prevalence of cardiovascular disease doubled from 22% (1992) to 41% (2012) and for ELSA 37% (2006) to 54% (2016). The prevalence of musculoskeletal disease (arthritis) was high at all data collection points, but there was no observable trend in cerebrovascular (stroke), endocrine (diabetes) and respiratory disease.

**Figure 2:**
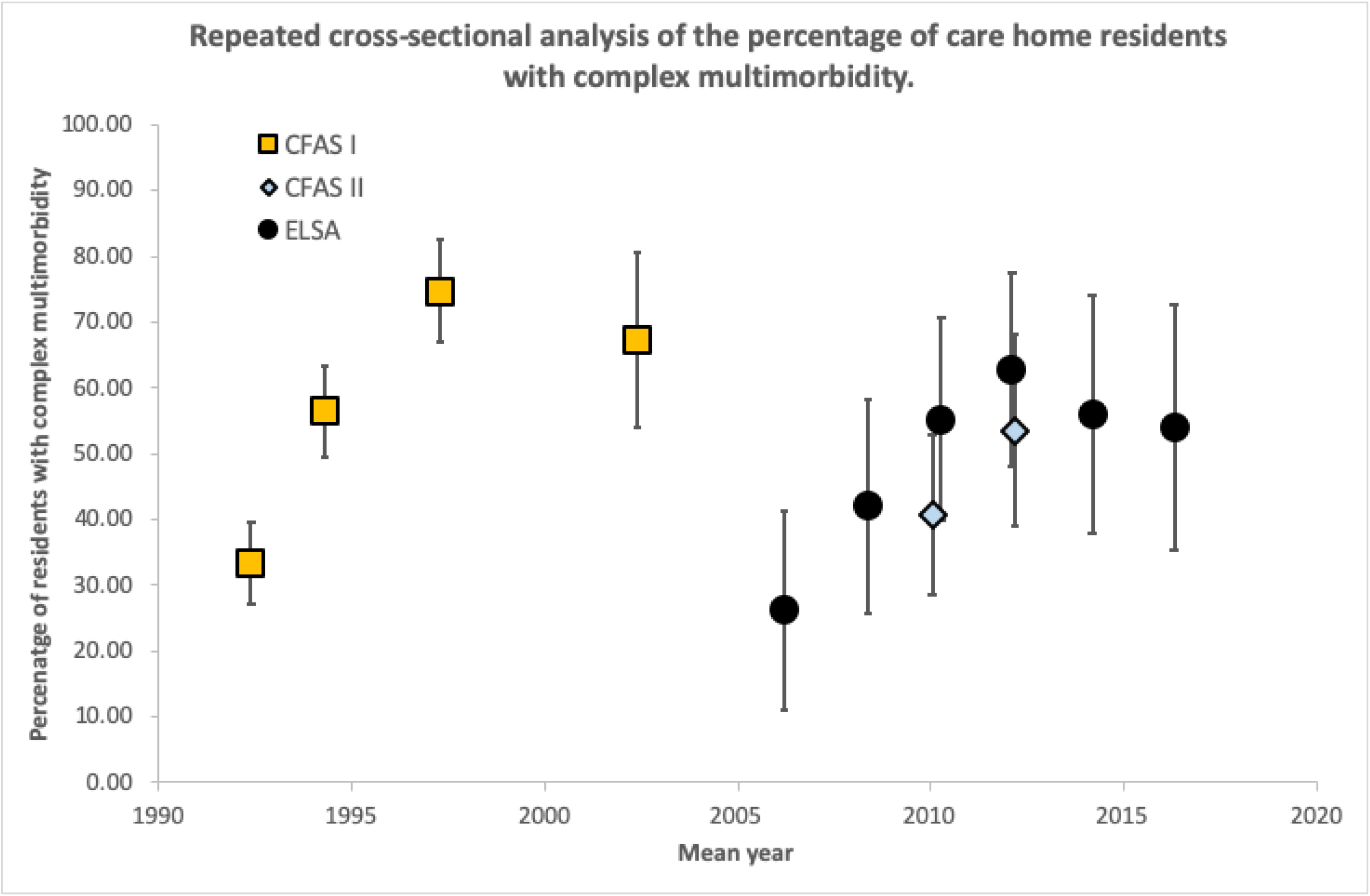
Prevalence of complex multimorbidity amongst care home residents from 1992-2016 in CFASI/II and ELSA studies. Complex comorbidity defined by medical conditions in at least three out of six domains (cardiovascular, cerebrovascular, musculoskeletal, respiratory, endocrine, cognition)

### Self-rated health

Figure 3 shows that self-rated health does not appear to change across the study period. It remained constant in CFAS I and CFAS II as nearly 50% of care home residents expressing fair/poor health. This was higher in ELSA, but with wide confidence intervals as informant interviews could not be used for this variable.

**Figure 3:**
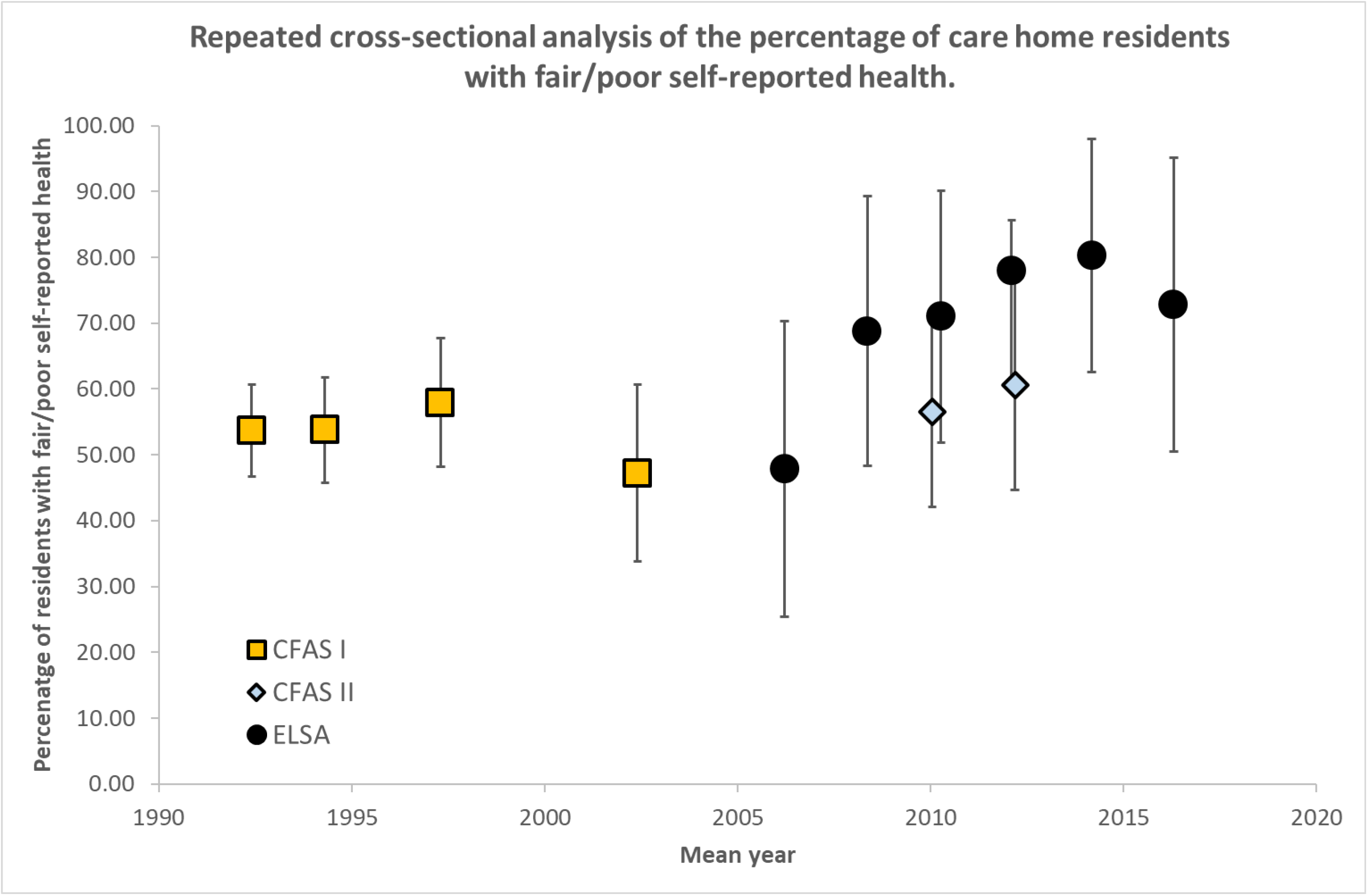
Prevalence of fair/poor self-reported health amongst care home residents from 1992-2016 in CFASI/II and ELSA studies.

### Sensitivity analysis

Similar results were seen if only responses from the first interview residents undertook in the care home setting were used to investigate the trends.

## Discussion

Over two decades, there has been an increase in the levels of disability amongst care home residents in England and Wales, as well as increases in complex multimorbidity demonstrated within two population-based cohort studies. Compared to residents in preceding decades, this study suggests that current care home residents have greater needs for support and consequently are likely to place greater demands on the staff and services who care for them.

### Comparison with other work

This study substantiates the perception that meeting the needs of care home residents is becoming more demanding and complex. It adds to current evidence indicating that, over time, older people in a range of settings are living with more medical conditions and more severe levels of disability [6,7,8]. The trends reported here were observed in both the CFAS and ELSA cohorts.

### Strengths and limitations

This is a novel, detailed exploration of how the health profile of UK care home residents has changed over more than two decades. A clear picture has emerged from analyses of data drawn from nationally representative studies that differed in their base populations, time, scope of data collection and purpose. This increases our confidence that the findings are representative of what is happening in care homes in England and Wales. However, it is important to acknowledge the limitations of our study design. None of the studies were designed to specifically investigate the health of care home residents, and some of the available measures of disability, such as ability to shop, were not relevant to this population. As expected, the variables in the two datasets were not identical. There were differences in how the questions were asked. CFAS participants were asked to distinguish between 'difficulty performing' and 'needing assistance' with ADLs, for example. To harmonise constructs such as disability or multimorbidity across studies, the study team was obliged to make some judgements on the comparability of measures. Nevertheless, despite these limitations, the message across different measures and datasets was the same.

### Implications for future research

Our findings emphasise the importance of developing systematic data collection on the health and functioning of care home residents in the UK [3], similar to the minimum dataset in the United States [13]. If data on the health of care homes residents had been available in the early stages of the COVID-19 pandemic, it could have informed management in care homes and may have influenced outcomes. In the longer term, access to a standardised set of information about the health and function of care home residents would allow more detailed exploration of changes in health and functioning for care home residents, without the need for resource intensive cohort studies.

As the complexity of residents' needs intensifies, the number of people in care homes is also expected to increase. The size of the care home resident population aged over 65 years has not changed since 2001, despite an 11% increase in the overall population at this age [14]. However, the proportion of people aged over 85 living in long-term care establishments is increasing [15]. Future expected gains in later life expectancy will lead to substantial increases in care needs, and it is predicted that 71,125 more care home places will be required in the UK by 2025 to meet these demands [16]. The increase in care needs requires urgent attention from policy makers and care providers to secure the resources required for future care home residents, especially considering the impact of the COVID-19 pandemic in care homes.

### Conclusion

This study demonstrates increases in the level of disability and the complexity of health problems amongst care home residents in England and Wales over two decades, which place additional demands on care home staff and health professionals. This should be an important concern for policymakers and service commissioners to ensure that the complex needs of care home residents are met in future, and when considering the impact of current and future waves of COVID-19 infection in this setting.

## Data Availability

Data were obtained from the Cognitive Function and Ageing Studies (MRC-CFAS CFAS I, CFAS II) and the English Longitudinal Study of Ageing (ELSA).

https://www.elsa-project.ac.uk/

http://www.cfas.ac.uk/

## Ethical approval

This study was approved by a University Research Ethics Committee (ref. 3791/2020).

